# Clinically validated metatranscriptomic analysis of human and microbial components of FFPE tissue biopsies

**DOI:** 10.1101/2023.05.03.23289449

**Authors:** Ryan Toma, Lan Hu, Diana Demusaj, Mory Mehrtash, Robert Wohlman, Guru Banavar, Momchilo Vuyisich

**Author notes:** Corresponding author: Ryan Toma.

## Abstract

Recent studies have identified microbial components in most tumors and invoked microorganisms in cancer onset and progression. The microbial functions responsible for these effects likely include production of genotoxins, modification of human gene expression, and protection of cancer cells from immune surveillance. Metatranscriptomics (MT) is a powerful tool for the development of potential cancer diagnostics and therapeutics, as well as investigating cancer initiation and progression. This is because MT data can quantify human and microbial gene expression, as well as provide strain level taxonomic classification of the microorganisms in the tumor microenvironment (TME). In addition, the microbial data derived from the MT method can easily be normalized across different samples, since many human transcripts can act as internal standards. When collecting tissue samples for clinical studies, researchers have the option of using fresh or Formalin-Fixed Paraffin-Embedded (FFPE) samples. FFPE samples are much easier to study, due to their widespread availability and ease of collection, storage, and handling. Little research exists comparing FFPE samples and fresh tissues, and there is no literature examining the differences in microbial signatures between these two sample preservation methods. In this study, we analyzed matched FFPE samples and fresh tissue samples from colon polyps of 13 participants for microbial and human components. We found that our clinically validated MT method generated equivalent data from FFPE samples and fresh tissue samples with high concordance for human gene expression (Pearson 0.70), microbial species (Pearson 0.94), and KEGG Orthologs (Pearson 0.97). These data indicate that FFPE samples are suitable for use in metatranscriptomic analyses, which will enable more diverse and larger oncology studies, as well as any other studies that benefit from MT analysis of tissue samples.

## Introduction

Metatranscriptomic (MT) analysis is a quantitative molecular method that can be used to gain valuable insights into the onset and progression of chronic diseases and cancers in human tissue samples ^1 –4^. By studying the transcriptome, which includes messenger RNA (mRNA) and non-coding RNA, researchers can obtain a comprehensive understanding of the functional genes and pathways that are activated or suppressed in cancer cells ^4^. This information is vital for the development of more effective treatments and therapies, as well as for the identification of potential biomarkers for cancer diagnosis and prognosis ^4^. In addition to investigations of human gene expression in cancers, MT analyses also provide information about the tumor microenvironment (TME) and the potential role of microorganisms in cancer development and progression ^5–7^. Importantly, by leveraging functional analyses, metatranscriptomics is better able to interrogate microbiomes and their roles in disease than traditional metagenomic approaches ^8^.

Metatranscriptomics has already proven to be a valuable technique for gaining insight into cancers. For example, research from our laboratory has shown that MT analyses of liquid saliva biopsies can be used for highly accurate diagnostic tests for oral and throat cancers, utilizing both microbial and human molecular features ^3^. In addition, a plethora of research has been performed on cancer tissues showing how human gene expression profiles change between cancerous and healthy tissue ^9,10^. These insights have led to the development of numerous potential diagnostics and therapeutics ^11^. For example, the gene expression profile of 90 different genes can be used to accurately identify the primary site in Cancer of Unknown Primary (CUP), which can be used for targeted therapy options ^12^. A panel of 23 differentially expressed human genes have been used in the generation of highly accurate breast cancer diagnostic methods which could increase diagnostic accuracy ^13^. Human gene expression profiles have also been used to predict the effect of cancer treatment which could be used to better inform therapeutic approaches to cancer therapies ^14^.

In addition to elucidating the impact of human genes in cancer, MT can also uncover the roles of microorganisms in cancer. Recent research has shown that different microorganisms are involved in the initiation and progression of several different cancers ^15^. For example, *Helicobacter pylori* has been shown to cause the onset of gastric cancer via activation of host cell proliferation pathways through CagA and VacA virulence factors ^16^, *Fusobacterium nucleatum* has been shown to enhance colorectal cancer metastasis through human m6A dysregulation ^17^, and *Porphyromonas gingivalis* has been found to be involved in prostate cancer immune evasion via the upregulation of PD-L1, which results in heightened cancer progression ^18^. MT represents one of the most comprehensive molecular biology techniques for investigating cancers as it is able to analyze both human and microbial gene expression patterns, identifying distinct differences between healthy and cancerous tissues, which can ultimately be used in diagnostics and therapeutics. Moreover, MT can be used for strain level taxonomic classification of all viable microorganisms within the TME. Not only is MT quantitative, its output can accurately determine the abundance of microbial components relative to the human tissue.

Collecting human tissue samples for molecular analysis is a critical aspect of cancer research. Tissue samples can either be obtained fresh and immediately frozen or preserved after surgical removal, or as Formalin-Fixed Paraffin-Embedded (FFPE) samples, which has been preserved for indefinite ambient temperature storage. From a practical perspective, it is much easier to obtain FFPE samples compared to fresh tissue ^19,20^. Fresh tissue requires immediate preservation and requires deviation from the standard of care in most clinics, whose standard operating procedure is to collect FFPE samples, not fresh tissue. FFPE samples can be easily obtained from biopsy specimens and stored for extended periods of time, making it an accessible option for researchers who wish to study cancer onset and progression, both prospectively and retrospectively.

The comparison between FFPE samples and fresh tissue samples has not been well explored in the context of MT analyses. Previous research has shown that human RNA features from a wide variety of tissue types are comparable between FFPE samples and fresh frozen tissue samples ^19–22^. These studies suggest that the human RNA profiles of FFPE samples and fresh tissue are comparable. However, this research is limited and does not investigate the microbial signatures between the tissue types. Furthermore, to the best of our knowledge, there are no publications specifically addressing this shortcoming, thus warranting more research in the field.

The purpose of this study was to assess the viability of FFPE samples, compared with fresh tissue for MT analysis. We analyzed fresh tissue and FFPE samples from the same colon polyp biopsy of 13 participants and compared the two sample types for both human and microbial features. We demonstrate that FFPE is a suitable option for the MT analysis of tissue and that it compares well to fresh tissue for quantitative human gene expression, microbial taxonomic classification, and microbial gene expression.

## Methods

### Ethical conduct of research

This study (clinicaltrials.gov registration number NCT05368688) was conducted with a protocol and consent forms approved by an Institutional Review Board (IRB) accredited by the United States Health and Human Services. All participants consented to participate in the study.

### Method validation of fresh tissue samples

For the purposes of the method validation, fresh frozen human bladder, prostate, uterus, and breast tissues were obtained from BioIVT, preserved, and homogenized as described in the tissue sample collection section below. Each fresh tissue type originated from a unique donor. Triplicate technical replicates of the fresh tissue were processed and sequenced as described in the library preparation section below. One replicate was analyzed for RNA integrity via fragment analyzer (DNF-472-0500, Agilent). Pearson correlations were computed between technical replicates for each tissue type to determine the reproducibility of the test.

### Matched FFPE and Fresh Tissue Sample Collection

FFPE and matched fresh tissue samples were collected from 13 participants (supplemental table 1). Participant 228 provided only a fresh tissue sample. This donor was removed from the comparative analyses, as they did not have a matching FFPE sample, but their tissue data is reported in supplemental tables 3-14. FFPE and matched fresh colorectal tissue samples were collected from the same colon polyp (all polyps were 8mm or larger). Fresh tissue was excised first and either placed directly into our custom RNA Preservative Buffer (RPB) before being homogenized using the Covidien Precision™ Disposable Tissue Grinder Systems (06-434A, Fisher Scientific) or fresh frozen at -80C until preservation and homogenization as above (supplemental table 2). Most samples were preserved or frozen immediately; however some samples remained at room temperature for a maximum of two hours prior to freezing or preservation, as the hospital staff were unable to immediately preserve them. FFPE samples were collected from the polyp after removing the fresh tissue samples. FFPE samples were immediately preserved in 10% Neutral Buffered Formalin (NBF). FFPE samples were kept in 10% NBF for a maximum of 12 hours prior to fixation. Three 10 micron FFPE slices from the same FFPE block were then prepared and analyzed from eight participants and two 10 micron FFPE slices from the same block were prepared and analyzed from five participants.

All samples were stored at -80°C upon arrival at our laboratory. Samples were analyzed within an average of 70 days (standard deviation of 39 days) of collection.

### Library Preparation and Sequencing

For the isolation of nucleic acids from the tissue samples, commercial kits were used according to manufacturer instructions. Direct-zol RNA Miniprep was used for the extraction of fresh tissue (R2050, Zymo Research). Quick-DNA/RNA FFPE Miniprep was used for the extraction of FFPE samples (R1009, Zymo Research). Following RNA extraction, all downstream molecular biology steps were performed using our clinically validated metatranscriptomic method, as previously published ^23^. The method includes DNase treatment, non-informative RNA depletion, cDNA synthesis, size selection, and limited cycles of PCR for adding dual unique barcodes to each sample. All samples were sequenced using the Illumina NovaSeq 6000 platform with 2x150 paired-end read chemistry.

### Bioinformatics

#### Microbiome

Our laboratory maintains a custom reference catalog which includes 32,599 genomes from NCBI RefSeq release 205 ‘complete genome’ category, 4,644 representative human gut genomes of UHGG^24^, ribosomal RNA (rRNA) gene sequences, and the human genome GRCh38^25^. These genomes cover archaea, bacteria, fungi, protozoa, phages, viruses, and the human host. The microbial genomes have 98,527,909 total annotated genes. Our laboratory adopts KEGG Orthology (KO) ^26^ to annotate the microbial gene functions using eggNOG-mapper^27^.

The microbiome pipeline maps paired-end reads to this catalog using Centrifuge^28^ for taxonomy classification (at any taxonomy rank). Reads mapped to the host genome and rRNA sequences are tracked for monitoring but excluded from further analysis. Reads mapped to microbial genomes are processed with an Expectation-Maximization (EM) algorithm^29^ to estimate the expression level (or activity) in the sample. Respective taxonomy ranks (strains, species, genus, etc.) can be easily aggregated from the genomes. For this study, we use species activity in the downstream analyses. These genome mapped reads are extracted and mapped to only gene or open reading frame (ORF) regions for molecular function or KO annotation and quantification.

The read counts of identified species and KOs are normalized by the total mapped reads in the sample in Count Per Million (CPM). We define the number of reads mapped to the microbiome as ‘microbial ESD’ (Effective Sequence Depth) to represent the usable portion of reads in a sample for microbiome.

#### Human transcriptome

A human transcriptome catalog is created based on the transcripts derived from Ensembl release 107^30^ annotations for GRCh38^25^ with transcripts missing in Ensembl but present in RefSeq ^31^. Paired-end reads are mapped to this transcript catalog and quantified with Salmon^32^. Human gene richness is the number of unique human genes, excluding the genes targeted for depletion, to which at least one read pair aligned. Transcript expression levels are then aggregated by gene to obtain gene expression levels. The read counts of genes are normalized to the total mapped reads excluding the probe targeted genes in CPM.

All data analysis and statistical tests were performed in R.

#### Quality Control

Samples had to pass quality control to be included in downstream analyses. Samples needed to have at least 6,000,000 reads mapped to the human transcriptome. Two FFPE samples were removed from the analyses (1CR418137322_1 and 1CR584970841_1) as they had less than 6,000,000 reads mapped to the human transcriptome. For the analysis of matched FFPE samples and fresh tissue, six external Contamination Controls (CCs) were analyzed on the same batch as the samples. CCs contain trace amounts (0.1pg) of synthetic RNA which is sufficient to produce a sequencing library and detect low levels of contaminants. Any microorganisms detected in the CCs were then removed from the FFPE and fresh tissue samples. On average, 14 species were removed from each FFPE sample and fresh tissue sample (standard deviation: 6).

## Results

### Method validation using fresh frozen tissue samples

To validate the MT analysis of fresh frozen tissue samples, technical replicates of fresh frozen tissue were analyzed. Fresh frozen tissue samples showed an average RNA Integrity Number (RIN) of 4.7 with a range of 3.6-6.9. While no current literature replicates our laboratory’s method for the preservation and homogenization of fresh tissue, the RNA integrity of the fresh tissue samples were similar to previously published literature ^20,33^, showing that the method is able to isolate RNA that is suitable for downstream MT analyses^34^.

Sequencing metrics reveal adequate sequencing quality for human genes, with an average of 12,825 human genes detected. Species richness was also adequate with an average of 27 species detected per sample. These results are roughly similar to previously published literature ^5,19,20^. No literature could be found examining the KO richness of fresh tissue samples; however, the low KO richness (on average 155 KOs) is expected due to the low species richness in tissue samples. Importantly, Pearson correlations between fresh tissue technical replicates show high concordance for human gene expression (average Pearson correlation: 0.975, standard deviation: 0.02), microbial species (average Pearson correlation: 0.983, standard deviation: 0.02), and KOs (average Pearson correlation: 0.890, standard deviation: 0.14).

These results indicate that our MT method generates reproducible results from fresh frozen tissue samples, with metrics that are similar to previously published methods.

### RNA yield and sequencing metrics on FFPE and fresh tissue samples

On average 1,382ng of RNA was recovered from FFPE samples and 2,273ng of RNA was recovered from fresh tissue samples. On average the FFPE samples received 221,557,685 total single reads and 350,504 microbial ESD (standard deviation of 56,981,468 total single reads and 192,561 microbial ESD). On average the fresh tissue samples received 233,052,824 total single reads and 405,930 microbial ESD (standard deviation of 49,551,349 reads and 164,916 microbial ESD).

FFPE samples detected a similar number of species, KOs, and had a similar percentage of total reads aligning to our custom reference catalog (%Microbial ESD) compared to fresh tissue (p>0.05, Figure 1A, B, and C); in FFPE samples, reads aligned to more human genes than the fresh tissue (p=0.0099, Figure 1D).

**Figure 1:**
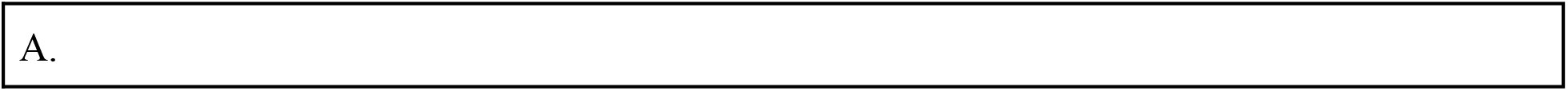
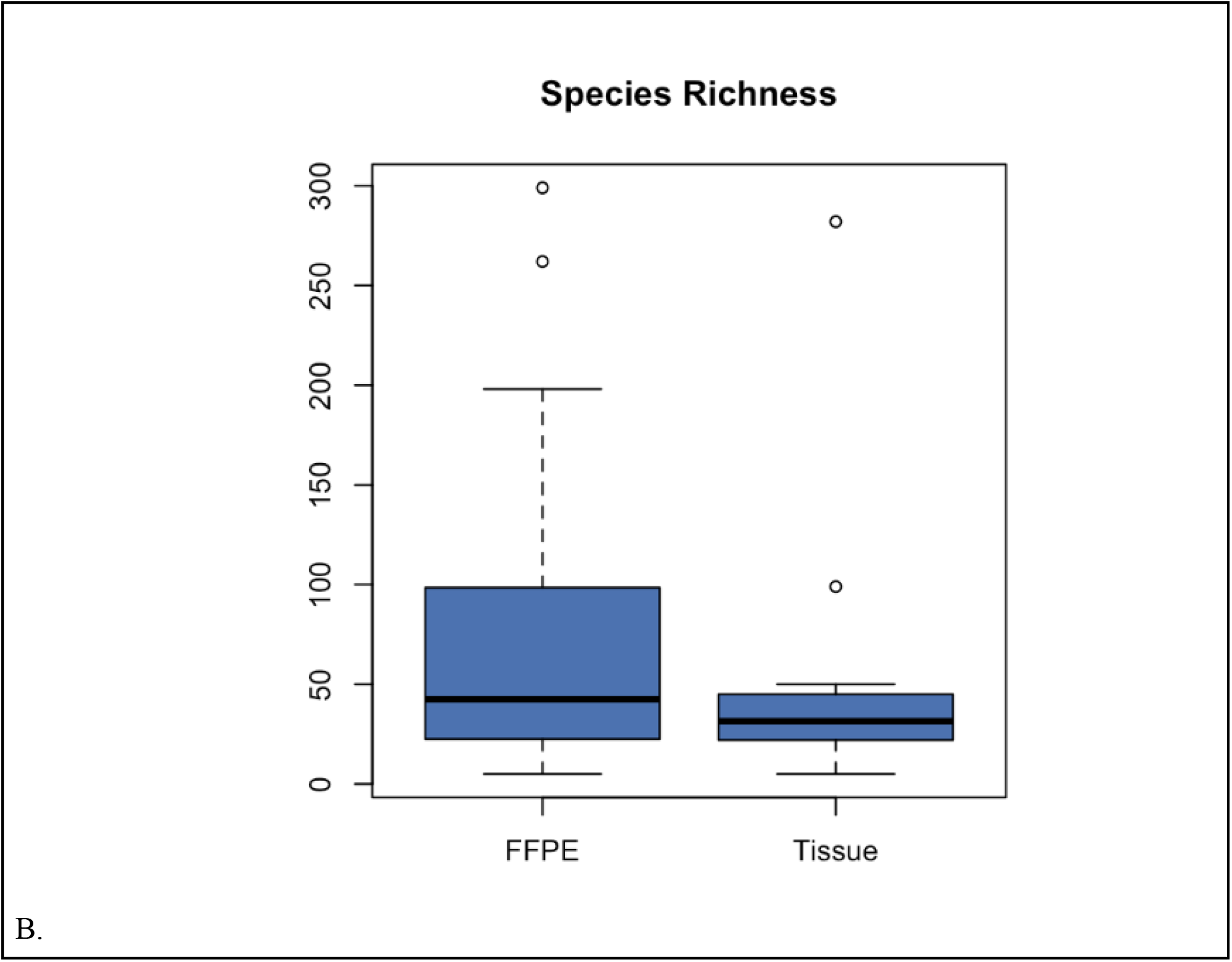
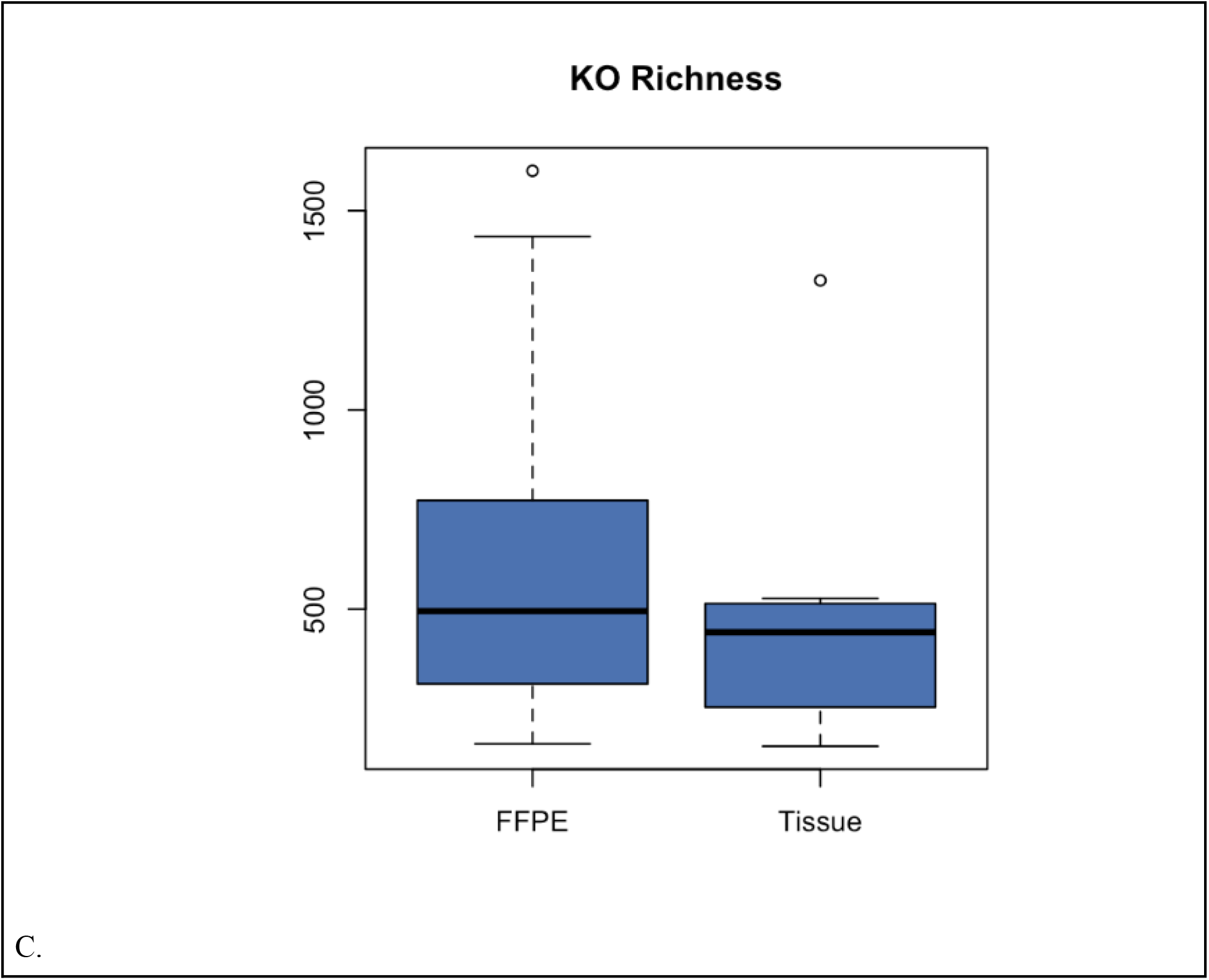
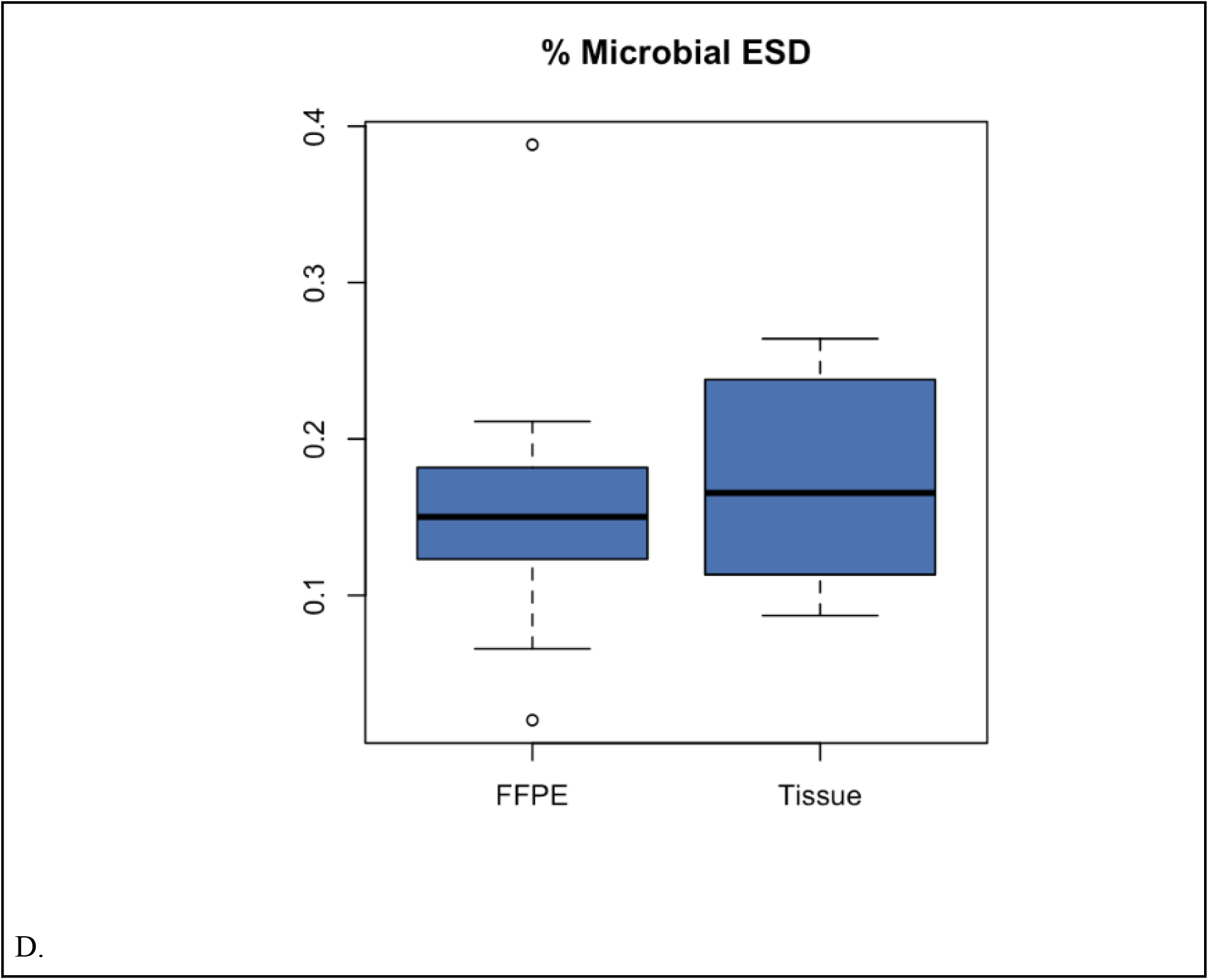
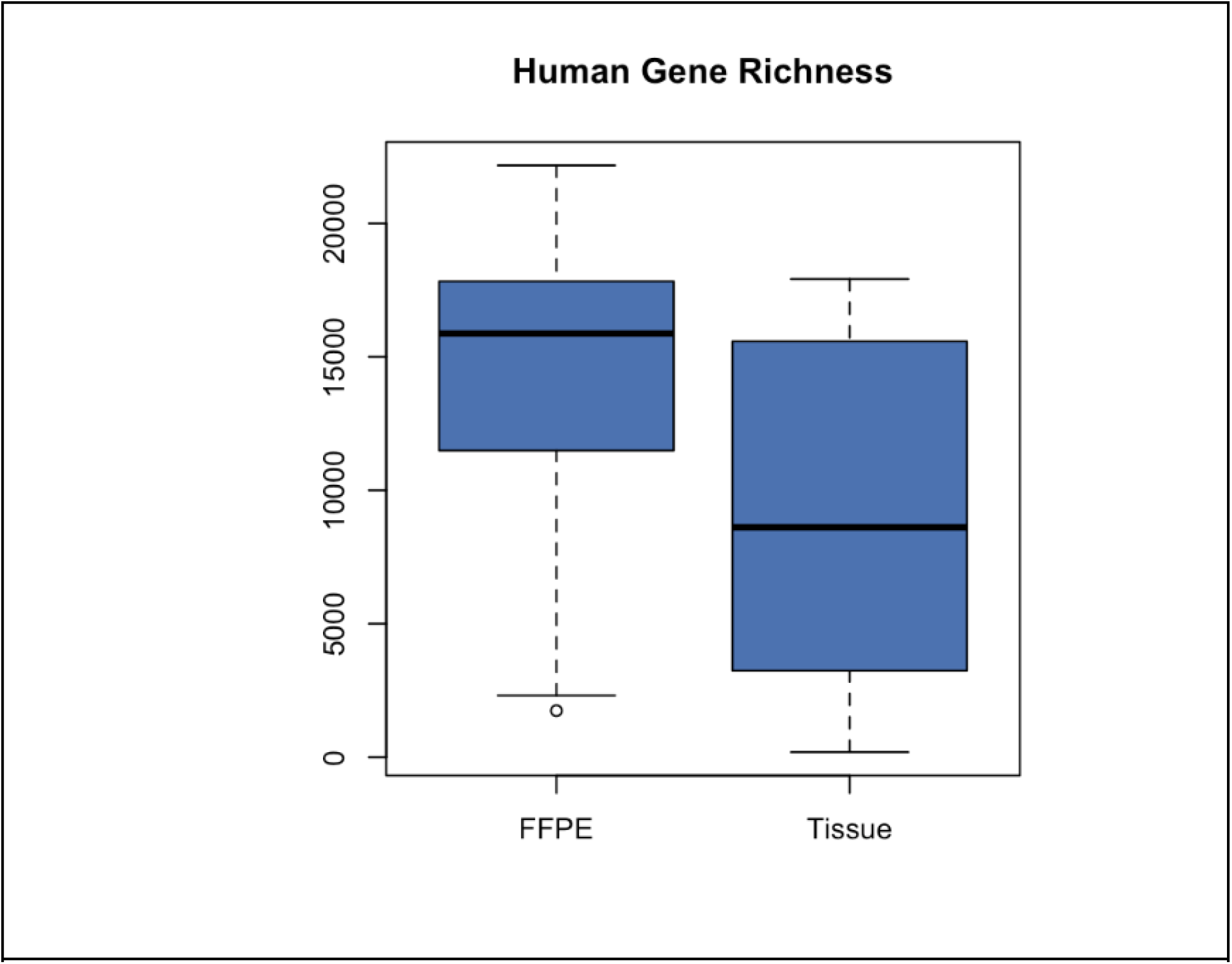
FFPE samples perform comparably or better to fresh tissue for basic sequencing metrics. FFPE samples detected a similar number of species (A), KOs (B), and had a similar %Microbial ESD (C) compared to fresh tissue (p>0.05); FFPE samples detected more human genes (D) than the fresh tissue (p=0.0099).

These results indicate that ample RNA can be obtained from both FFPE and fresh tissue samples for MT analysis. FFPE samples also compare well to fresh tissue samples on basic sequencing metrics with an improvement in the number of human genes to which reads could be mapped. For a full list of the relative abundance of all species, KOs, human genes, and their read counts in each FFPE and fresh tissue sample see supplementary tables 3-14.

### Correlations between matched FFPE and fresh tissue samples

FFPE samples correlate well with fresh tissue samples for KOs (average Pearson correlation: 0.965; standard deviation: 0.036; Figure 2), microbial species (average Pearson correlation: 0.939; standard deviation: 0.086; Figure 2), and human gene expression (average Pearson correlation: 0.696; standard deviation: 0.139; Figure 2).

**Figure 2:**
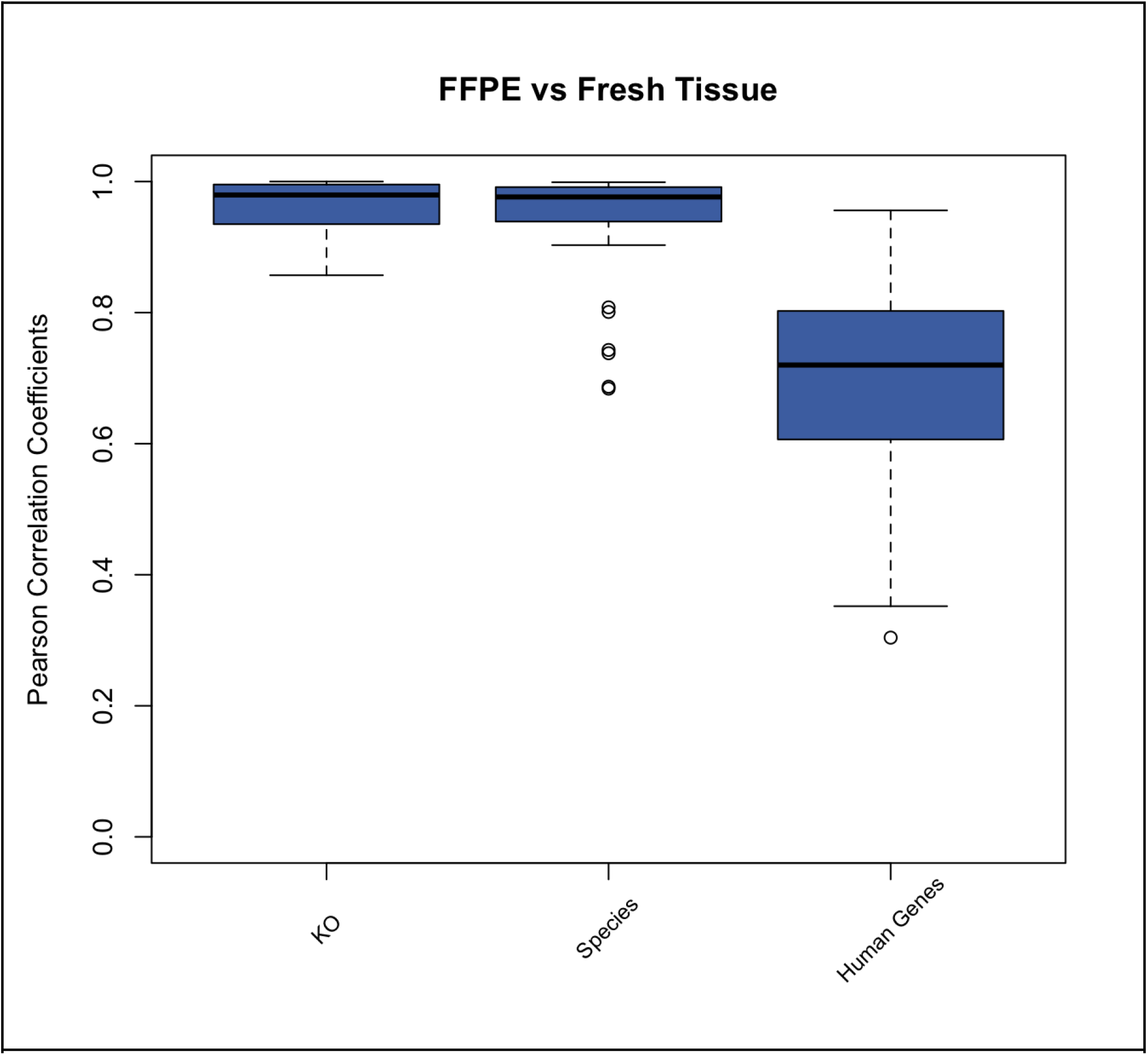
FFPE samples correlate well with fresh tissue samples for KOs, microbial species, and expression of human genes.

These results demonstrate that FFPE samples maintain the distribution of microbial and human signatures seen in fresh tissue samples.

### Method precision for the analysis of FFPE samples

FFPE sample replicates from the same donor correlate well with one another for KOs (average Pearson correlation of 0.99; standard deviation: 0.021; Figure 3), microbial species (average Pearson correlation of 0.969 standard deviation: 0.045; Figure 3), and human gene expression (average Pearson correlation of 0.902; standard deviation: 0.12; Figure 3).

**Figure 3:**
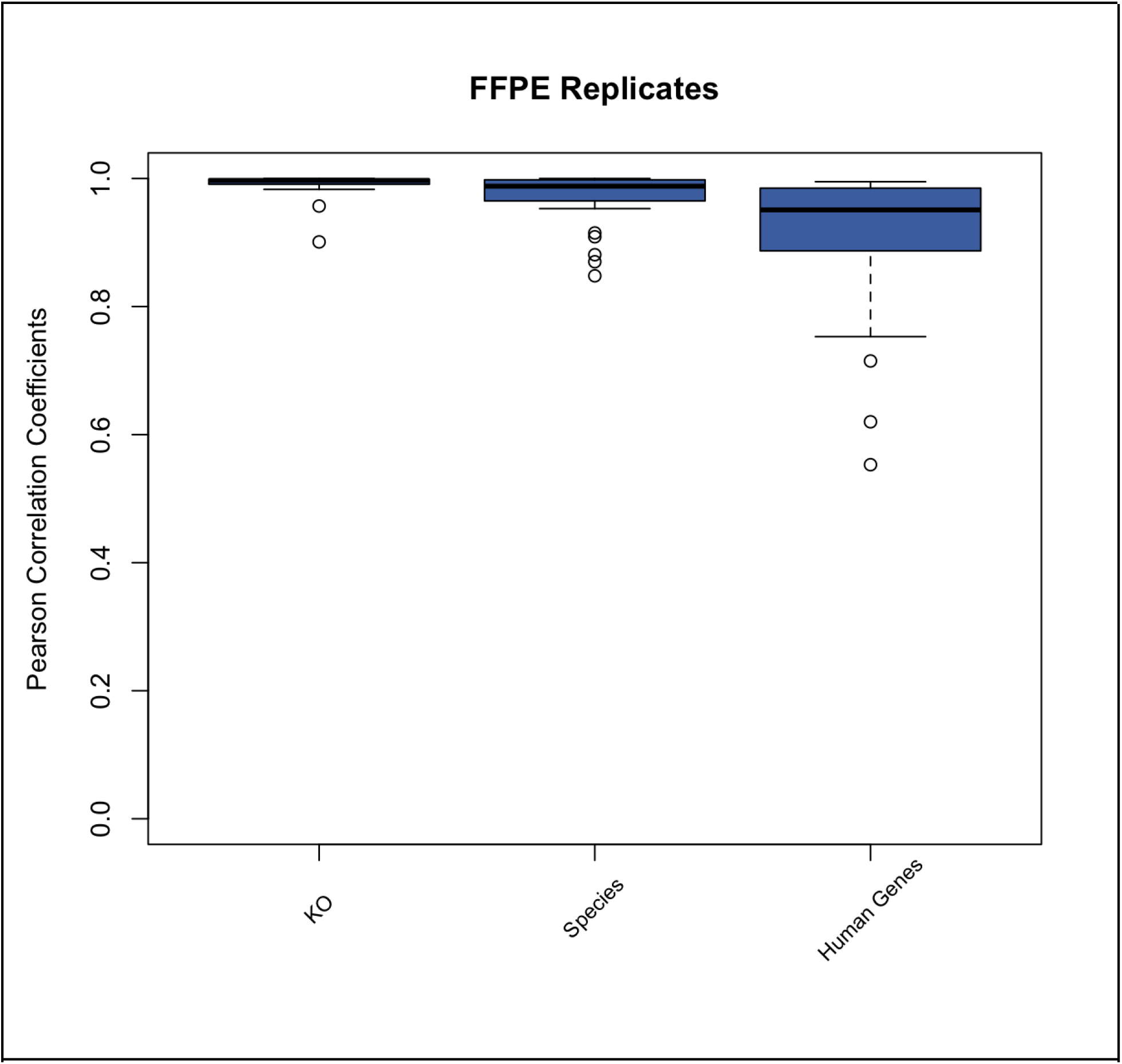
Biological replicates of FFPE samples correlate well. FFPE sample replicates from the same donor correlate well with one another for KOs, microbial species, and human gene expression.

These results show that the method used for the MT analysis of FFPE samples is highly reproducible.

## Discussion

The analysis of tissue biopsy samples is critical for developing an understanding of cancer initiation and progression. In particular, MT investigations of cancer tissue have proven to be invaluable, elucidating both the role of microorganisms and human genes ^1 –5,9,10,12 –18^. However, one of the main difficulties in analyzing tissue samples is obtaining fresh tissue ^35^. Fresh tissue is not collected by healthcare providers as part of the standard of care and modifying their existing standard operating procedures is difficult. Therefore acquiring fresh tissue samples for analysis is challenging, thus limiting the studies that include tissue biopsies. Meanwhile, FFPE samples are readily available. In addition to their ubiquitous collection, FFPE samples are also routinely archived, enabling large scale retrospective studies.

Herein we analyzed matched fresh and FFPE samples from 13 participants with MT. We showcase that the methodology utilized by our laboratory for the analysis of both FFPE and fresh tissue samples is able to isolate adequate amounts of RNA, is reproducible, and can generate informative sequencing data. Figure 1 shows that FFPE samples perform better than fresh tissue for the detection of human genes, while performing comparably to fresh tissue samples for all other critical sequencing metrics (species richness, KO richness, and %Microbial ESD). We corroborated previous literature showing that human gene expression profiles are highly conserved between tissue types (figure 2), indicating that in the context of MT, FFPE samples are a suitable sample type for the investigation of human gene expression profiles ^19,20,35^. Importantly, to the best of our knowledge, this is the first paper to also show that microbial gene expression and microbial species taxonomic classification are well correlated between FFPE samples and fresh tissue (figure 2). This finding is critical as there has been increasing research showing the importance of the tumor microbiome and microbial molecular features in cancer initiation and progression. Our findings showcase that FFPE samples are suitable for interrogating these features with MT^36–40^.

A potential limitation of the present study was that the FFPE samples were analyzed 70 days after collection, so we are unsure of the applicability of the results to retrospective studies that are analyzing FFPE samples that have been in storage for much longer periods of time. Future studies should be performed to gain a robust understanding of how RNA sequencing data from FFPE samples are affected by long-term storage and how this relates to degradation observed in fresh tissue. An additional limitation of this study is the length of time that the fresh tissue remained at room temperature prior to preservation (<2 hours). The clinic was not able to adjust their procedures to always preserve the fresh tissue immediately, which is a limitation to the adoption of fresh tissue collection in clinics as modifications to standard operating procedures are often not possible. The amount of time the fresh tissue remained at room temperature could have reduced data quality and altered the correlations to the FFPE samples. Further research should be conducted on immediately preserved tissue versus FFPE samples to confirm the results of this study.

MT is a valuable tool that can be used to investigate cancer initiation and progression as well as develop potential diagnostics and therapeutics. Logistically, analyzing FFPE samples is preferred over fresh tissue due to the ease of collection and its widespread accessibility. By using MT we demonstrated that FFPE tissue is comparable or better than fresh tissue for sequencing metrics, human and microbial gene expression profiles, and microbial taxonomic classification. FFPE tissue samples can and should be used for prospective and retrospective MT research.

## Data Availability

All data produced in the present study are proprietary to Viome Life Sciences but may be available upon reasonable request to the authors

## Author Contributions

MV, MM, and RW designed and implemented the study. RW and MM performed metadata and sample collection. RT developed the laboratory analysis methods and performed the molecular analyses for the matched FFPE and tissue samples. DD performed the molecular analyses for the fresh frozen method validation. LH and GB performed data analysis. LH, MV, and RT contributed to data interpretation. All authors contributed to the writing of the manuscript.

